# Peri- and Post-natal Risk Factors Associated with Health of Newborns

**DOI:** 10.1101/2023.01.12.23284503

**Authors:** Yanan Long, Atif Khan, Andrey Rzhetsky

**Affiliations:** Department of Chemistry, The University of Chicago, 900 E. 57^th^ Street, Chicago, 60637, IL, USA; Department of Medicine, The University of Chicago, 900 E. 57t^h^ Street, Chicago, 60637, IL, USA; Department of Human Genetics and Committee on Quantitative Methods in Social, Behavioral, and Health Sciences, The University of Chicago, 900 E. 57 Street, Chicago, 60637, IL, USA

**Keywords:** Newborn immunity, maternal factors affecting child health, susceptibility to infections, immune system disbalance risk factors

## Abstract

Designing prophylactic strategies for newborns requires understanding of the factors that contribute to immunity and resistance to infection. We analyzed 1,892,035 mother-newborn pairs in which both the mother and newborn were observed continuously for at least one year before and after birth. As part of this study, we considered maternal exposures to infections and immune disorders during pregnancy, exposures to anti-infection medications by both mother and newborn, as well as the newborn’s delivery type and reported complications. According to our analyses, infection rates and immune disorder rates were over-dispersed among newborns. The most consequential factors predicting newborns’ immune health were preterm birth, with 276.3% and 193.9% risk increases for newborn bacterial infections. Newborn anti-infective prescriptions were associated with considerable increases in risk of diseases affecting immune health, while maternal prescriptions were associated with fewer outcomes and with mixed signs. The Cesarean section mode of delivery, the mother’s age, the sex of the newborn, and the mother’s exposure to infections all showed significant but smaller effects on the newborn’s immune health. Female newborn appeared to be better protected against diseases with immune system etiology, except for miscellaneous infections.

---

Accumulating evidence suggests that a mother’s microbiome composition, along with that of her newborn child are consequential for the child’s long-term predisposition to a range of diseases, ^1,2^ including infections, immune-related diseases,^3^ allergies, ^4^ and malignancies.^5^ A somewhat contested topic around factors affecting child health is how a Cesarian-section vs. vaginal delivery, or preterm birth, will affect the newborn’s long-term health. ^5^ While antibiotics and preventive vaccinations have reduced infectious disease prevalence worldwide, pediatric immune system disorders have increased concurrently.^6^

In this study, we used a very large observational dataset to investigate peri- and postnatal factors putatively affecting newborns.^7^ These factors included infections affecting pregnant mothers and newborns, the mode and term of delivery,^8^ and anti-infective medication use by pregnant mothers and the newborns. ^9^

For our analyses, we used the MarketScan commercial insurance dataset, covering health events of more than half of the US population during 2003–2018.^10^ The data provide person- and daily-level resolution of disease diagnoses, prescription medication, medical procedures, and family linkage information inferred from co-insurance data. Our analysis of these data enabled us to calculate mother-child links for over 3.5 million mother-child pairs, of which 1,892,035 were “visible” for at least a year before and after the live births. ^11,12^ We matched newborn babies with their mothers using the following definitions: A newborn was identified as a new MarketScan enrollee with an appropriate age (0 years) and at least one live-birth diagnostic code in the International Classification of Diseases (ICD, versions 9 and 10). ^13,14^ We identified a mother as a pregnant woman covered by the same family insurance policy as the newborn, and a mother was also identified as having at least one live-delivery-related diagnostic code registered within three days of the first code.

We analyzed a range of regression models and performed a comprehensive model selection (see *Supplement* 1.3.3), and present here the results for the models that best fit the data for each outcome phenotype. For each of the outcome phenotypes, the full model (with all predictors) was the optimal. The outcome variable in each of our regression models was an integer representing the counts of events (such as bacterial or viral infections), in a child’s life. Had our cohort been homogeneous, in which the rate of a rare event was the same for each person, we would expect that health event counts would follow a Poisson distribution. Instead, our model selection analysis shows a disease event distribution variance over newborns that is much greater than the mean of the corresponding distribution.

Therefore, we further assumed that the cohort outcomes were generated by a mixture of Poisson processes, each with rate *λx*, where *λ* is the overall mean rate across the whole cohort, and *x* is a random variable with an expected value of 1, sampled from a Γ-distributed random variable with shape parameter *r* and mean 1, (cf. Figure 1, plates (B) and (C)). This convolution of Γ- and Poisson distributions results in a negative binomial (NB) distribution of outcome counts. As parameter *r* tends to zero, Γ-distribution tends to a *δ*-function, and the overall count distribution across the cohort tends to a Poisson distribution. Our estimated values of *r* were less than one for all four outcome events, suggesting that the distribution of health event rates is L-shaped. This L-shaped distribution (see Plate (C) in Figure 1) indicates that rates of health events varied across individuals in the cohort, with the distribution’s heavy right tail describing extremely high event rates. To flexibly model the counts of zero-counts in the outcomes, we also used hurdle NB models. Model comparison showed that the hurdle NB model was the best fit for all but miscellaneous infections, for which the (full) hurdle and non -hurdle models were not significantly different from each other (see Supplementary Tables S5 & S6, Figure S4). We implemented our models using the `Stan` probabilistic programming language^15^ with the `cmdstanr` interface.^16^ For each model, we ran 16 independent Markov chain Monte Carlo (MCMC) processes with 250 warm-up and 250 sampling iterations with diffuse initialization. To assess model convergence, we checked the default metrics provided in `Stan` (see *Supplement* 1.3.3). For model comparison, we initially used the leave-one-out information criterion (LOOIC) approximated via Pareto smoothed importance sampling (PSIS ^17^), but then used 10-fold cross-validation (CV) due to large numbers of problematic data points. We used the packages `brms`,^18^ `loo`,^19^ `posterior`,^20^ and `bayesplot` for defining our models, computing information criteria values, posterior processing and visualization, respectively. We used the R programming language (version 4.1.1)^21^ for all our analyses.

**Figure 1.**
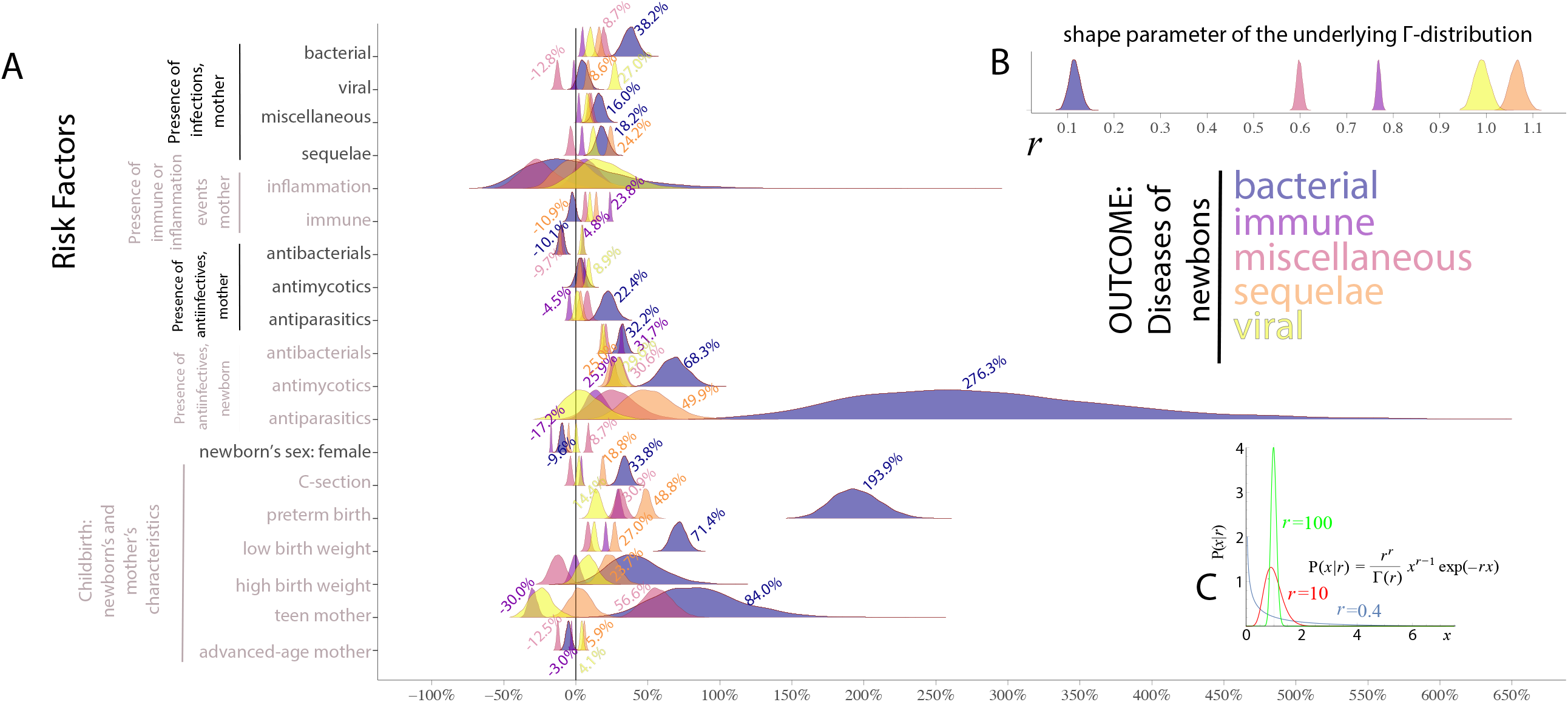
Estimating the effect of the maternal, fetal, and neonatal risk factors associated with infections and immune disorders in infants. Plate (A): Posterior distributions of estimated effect sizes for fetal and neonatal risk factors. The *Y* -axis shows labels for distinct disease risk factors, while the *X*-axis represents effect size in per cents. **Plates (B) and (C):** Γ**-distribution properties underlying newborn disease rate distribution**. (B) Γ-distribution shape parameter estimates describing disease rate variation among infants for several outcome conditions: bacterial infections (blue); miscellaneous (violet); sequelae (salmon); viral (yellow); and immune disorders (magenta). The scale parameter of the Γ-distribution is fixed, so that the distribution’s mean is always equal to one. According to the modeling formalism, disease counts for individual patients are sampled from a Poisson distribution with rate λ *x*, where random variable *x* is sampled from a Γ-distribution with mean 1 and shape parameter *r*. Γ-distribution shape parameter value *r <* 1 defines a *L*-shaped distribution, see Plate C. This asymmetric distribution suggests that most infants have a very limited response to risk factors, but a small proportion of (the distribution’s heavy right tail) is associated with a powerful response. The Γ-distribution tends to a Poisson distribution as the shape parameter value grows, see Plate C.

We report all results as risk changes in terms of relative expected counts (see *Supplement* 1.2) obtained using the best-fit model for each outcome phenotype, as determined by the model selection procedure, but in the text below, we only mention predictors with statistically significant associations with newborn health.

First, a wide range of anti-infective prescriptions given to newborns during the first six months of life and of anti-infective medications given the mothers prior to birth were associated with altered risks of immune system outcomes for the newborn. Newborn medications were associated with marked risk increases for immune system-related diseases. In particular, the presence of antiparasitic drugs was associated with a 276.3 percent and a 49.9 percent increase in risk of bacterial and sequelae infections, respectively. For antibacterial medications, the risk increases were 32.2 percent, 20.8 percent 19.2 percent and 18.1 percent for bacterial, miscellaneous, viral and sequelae infections, respectively; risk of immune disorders increased by 31.7 percent. Finally, antimycotic medications predicted risk increases of 68.3 percent, 30.6 percent, 29.6 percent, and 25.0 percent for bacterial, miscellaneous, viral and sequelae infections, respectively, whereas for immune disorders the risk increase was 25.9 percent.

Second, medications administered to the mother turned out to have rather diverse effects on her newborn’s immune health. Somewhat surprisingly, antibacterial medications appeared to lower risks for newborn’s bacterial-(-10.1 percent), sequelae-(-10.9) and miscellaneous (-9.7 percent) infections. The same medications were associated with risk increases for immune disorders (4.8 percent) and viral infections (4.3 percent). Antiparasitic medications were associated with risk increases of 22.4 percent and 7.9 percent for bacterial and miscellaneous infections, respectively, while the risk decreased by 4.5 percent for immune disorders. Finally, for antimycotics, the only significant result was viral infections (8.9 percent).

Third, we observed that the immune health of newborns in the cohort is extremely heterogeneous: counts of immunity-related disorders in the cohort were over-dispersed compared to the Poisson distribution of outcome events with the same mean. In other words, a distribution of rates of diseases per infant was L-shaped, with a long right tail of the distribution describing infants with the highest rates of diseases.

Fourth, we our results suggested that preterm birth, teenage pregnancy, and Cesarean delivery were the most consequential for newborn immune health. Female newborns had a lower risk of immune-related diseases than male, with an exception for miscellaneous infections, whereas teenage births had variable-sign effects on the outcomes:

- As expected,^22^ female newborns appear to be better protected against diseases with immune system etiology. We estimated a -9.6 percent risk change for bacterial infections in female newborns, a -5.0 percent for sequelae infections and a -17.2 percent for immune disorders. Only for miscellaneous infections did we observe a positive risk change for female newborns (8.7 percent).
- Preterm birth has strong associations with a child’s subsequent bacterial infections, with the disease risk increased by 193.9 percent. Similarly, this predictor was associated with 30.9 percent, 48.8 percent, and 14.4 percent risk increases of the child’s miscellaneous, sequelae, and viral infections, respectively. A child’s preterm birth is associated with a 29.3 percent risk increase of immune disorders.
- Childbirth via Cesarean section (C-section) was associated with 33.8 percent and 18.8 percent risk increases for newborn bacterial and sequelae infections, respectively. The effect size is not statistically different from zero for viral and miscellaneous infections. The risk increase associated with C-section is 3.8 percent for future immune disorders.
- Children with low birth weights (less than or equal to 2,500 g) had risk increases of 71.4 percent, 27.0 percent, 12.7 percent and 8.5 percent for bacterial, sequelae, viral and miscellaneous infections. The risk increase associated with low birth weights was 20.8 percent for immune disorders.
- Teenage pregnancy represents the most diverse effect-sign disease outcome, with a risk change of 84.0 percent and 56.5 percent for miscellaneous and viral infections, respectively. The effect size is not statistically different from zero for bacterial infections. The newborn having a teenage mother has the risk of developing immune disorders reduced by 30.0 percent.

Fourth, a pregnant woman’s history of infections and immune disorder predicts a predominantly significant, but smaller, magnitude change in her newborn’s disease risk, as shown in Figure 1 and Table 1. If a pregnant mother ever had bacterial infections, our study shows increases in risk of her newborn’s bacterial infections, miscellaneous infections, sequelae infections and viral infections of 38.2 percent, 19.8 percent, 16.4 percent and 10.4 percent, respectively. Presence of maternal sequelae infections during pregnancy predicts an increase in a newborn’s risk of the same kind of infections by 24.2 percent, and an increase of 18.2 in the risk of bacterial infections. Similarly, if a pregnant mother experienced bacterial infections, the newborn’s risk of contracting these infections increased by 38.2 percent. Finally, the presence of maternal viral infections is associated with a 27.0 percent increase in the risk of the same infections in the newborn. By contrast, the presence of maternal viral infections is associated with a -12.8 percent change in risk for newborn’s miscellaneous infections.

**Table 1.**
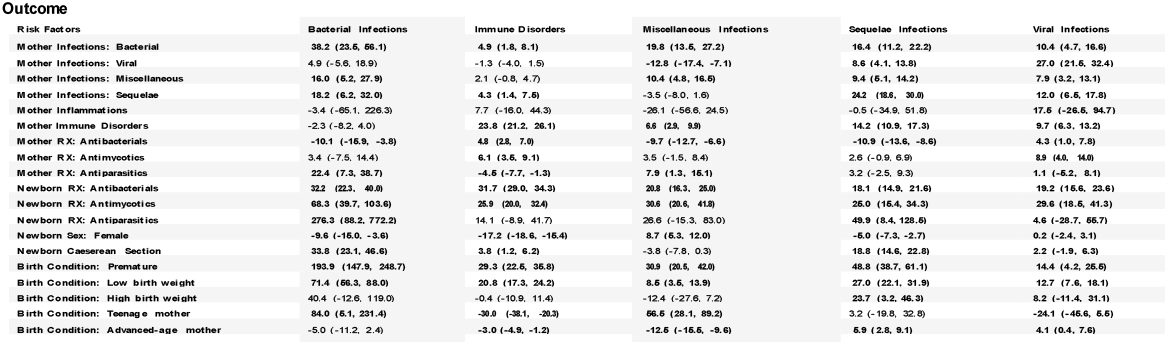
Effect size: posterior-probability estimates in per cents (%) associated with risk factors (posterior median) and 99.9 percent credible intervals (statistically significant results in **boldface**) for five outcomes of newborn health status: bacterial infections (intestinal infectious diseases, tuberculosis, zoonotic bacterial diseases, other bacterial diseases);, immune disorders (disorders involving the immune mechanism, such as nontoxic single thyroid nodule, diabetes mellitus, polyglandular dysfunction, amyloidosis, unspecified metabolic disorders, intracranial and intraspinal phlebitis and thrombophlebitis, multiple sclerosis, acute disseminated demyelination, demyelinating diseases of central nervous, myasthenia gravis and other myoneural disorders, hordeolum and chalazion, conjunctivitis, immune disorders of sclera, keratitis, iridocyclitis, chorioretinal inflammation, purulent endophthalmitis, rheumatic fever with heart involvement, rheumatic chorea, sinusitis, laryngitis, and epiglottitis, vasomotor and allergic rhinitis, Crohn’s disease, ulcerative colitis, noninfective gastroenteritis, irritable bowel syndrome, inflammatory liver diseases, lymphadenitis, pemphigus, dermatitis); miscellaneous infections (including mycoses, protozoal diseases, helminthiases, pediculosis, acariasis and other infestations), sequelae infections, and viral infections.

| Risk Factors |  | Bacterial Infections | Immune Disorders | Miscellaneous Infections | Sequelae Infections | Viral Infections |
| --- | --- | --- | --- | --- | --- | --- |
| Mother Infections: Bacterial |  | 38.2 (23.5, 56.1) | 4.9 (1.8, 8.1) | 19.8 (13.5, 27.2) | 16.4 (11.2, 22.2) | 10.4 (4.7, 16.6) |
| Mother Infections: Viral |  | 4.9 (-5.6, 18.9) | -1.3 (-4.0, 1.5) | -12.8 (-17.4, -7.1) | 8.6 (4.1, 13.8) | 27.0 (21.5, 32.4) |
| Mother Infections: Miscellaneous |  | 16.0 (5.2, 27.9) | 2.1 (-0.8, 4.7) | 10.4 (4.8, 16.5) | 9.4 (5.1, 14.2) | 7.9 (3.2, 13.1) |
| Mother Infections: Sequelae |  | 18.2 (6.2, 32.0) | 4.3 (1.4, 7.5) | -3.5 (-8.0, 1.6) | 24.2 (18.6, 30.0) | 12.0 (6.5, 17.8) |
| Mother Inflammations |  | -3.4 (-65.1, 226.3) | 7.7 (-16.0, 44.3) | -26.1 (-56.6, 24.5) | -0.5 (-34.9, 51.8) | 17.5 (-26.5, 94.7) |
| Mother Immune Disorders |  | -2.3 (-8.2, 4.0) | 23.8 (21.2, 26.1) | 6.6 (2.9, 9.9) | 14.2 (10.9, 17.3) | 9.7 (6.3, 13.2) |
| Mother RX: Antibacterials |  | -10.1 (-15.9, -3.8) | 4.8 (2.8, 7.0) | -9.7 (-12.7, -6.6) | -10.9 (-13.6, -8.6) | 4.3 (1.0, 7.8) |
| Mother RX: Antimycotics |  | 3.4 (-7.5, 14.4) | 6.1 (3.5, 9.1) | 3.5 (-1.5, 8.4) | 2.6 (-0.9, 6.9) | 8.9 (4.0, 14.0) |
| Mother RX: Antiparasitics |  | 22.4 (7.3, 38.7) | -4.5 (-7.7, -1.3) | 7.9 (1.3, 15.1) | 3.2 (-2.5, 9.3) | 1.1 (-5.2, 8.1) |
| Newborn RX: Antibacterials |  | 32.2 (22.3, 40.0) | 31.7 (29.0, 34.3) | 20.8 (16.3, 25.0) | 18.1 (14.9, 21.6) | 19.2 (15.6, 23.6) |
| Newborn RX: Antimycotics |  | 68.3 (39.7, 103.6) | 25.9 (20.0, 32.4) | 30.6 (20.6, 41.8) | 25.0 (15.4, 34.3) | 29.6 (18.5, 41.3) |
| Newborn RX: Antiparasitics |  | 276.3 (88.2, 772.2) | 14.1 (-8.9, 41.7) | 26.6 (-15.3, 83.0) | 49.9 (8.4, 128.5) | 4.6 (-28.7, 55.7) |
| Newborn Sex: Female |  | -9.6 (-15.0, -3.6) | -17.2 (-18.6, -15.4) | 8.7 (5.3, 12.0) | -5.0 (-7.3, -2.7) | 0.2 (-2.4, 3.1) |
| Newborn Caesarean Section |  | 33.8 (23.1, 46.6) | 3.8 (1.2, 6.2) | -3.8 (-7.8, 0.3) | 18.8 (14.6, 22.8) | 2.2 (-1.9, 6.3) |
| Birth Condition: Premature |  | 193.9 (147.9, 248.7) | 29.3 (22.5, 35.8) | 30.9 (20.5, 42.0) | 48.8 (38.7, 61.1) | 14.4 (4.2, 25.5) |
| Birth Condition: Low birth weight |  | 71.4 (56.3, 88.0) | 20.8 (17.3, 24.2) | 8.5 (3.5, 13.9) | 27.0 (22.1, 31.9) | 12.7 (7.6, 18.1) |
| Birth Condition: High birth weight |  | 40.4 (-12.6, 119.0) | -0.4 (-10.9, 11.4) | -12.4 (-27.6, 7.2) | 23.7 (3.2, 46.3) | 8.2 (-11.4, 31.1) |
| Birth Condition: Teenage mother |  | 84.0 (5.1, 231.4) | -30.0 (-38.1, -20.3) | 56.5 (28.1, 89.2) | 3.2 (-19.8, 32.8) | -24.1 (-45.6, 5.5) |
| Birth Condition: Advanced-age mother |  | -5.0 (-11.2, 2.4) | -3.0 (-4.9, -1.2) | -12.5 (-15.5, -9.6) | 5.9 (2.8, 9.1) | 4.1 (0.4, 7.6) |

Our study’s results resonate with a growing body of studies linking human microbiome states to immune system health.^23-25^ Our data suggests that dysbiosis, the disruption of healthy microbiome composition, is linked to a range of diseases,^26-28^ such as autoimmune, inflammatory diseases^29^ and infection responses.^27^ A plethora of prior studies suggests a putative causal scenario that might explain our findings (see Figure 2).

**Figure 2.** Putative causal relationship among factors contributing to the development of an infant’s immune system.

First, a preterm birth can change the development of a child’s innate and adaptive immunity,^30^ as well as alter interactions between the child’s immune system and microbiota.^31^ An analysis of premature newborns’ immune profiles demonstrated that they differ significantly from those of full-term babies.^32^ Similarly, a comparison of the diversity of newborns’ gastrointestinal microbiota^33^ has shown that preterm newborns had lower microbiota diversity than their full-term peers.

Second, the Cesarean delivery mode is associated with higher risks of immune system disorders later in life. For example, studies have shown a risk increase (from ten to 40 percent) in the immune system disorders of those Cesarean-born, such as immune deficiencies, asthma, and inflammatory bowel disease (IBD).^8,34^ The mechanism of this association is still disputed. One suggested explanation is that vaginal birth is critical for the transfer of a mother’s microbiome to her baby. However, a recent experimental study^35^ suggested that neonatal microbiome profiles are very similar between newborns delivered via the two different methods. Note that the same newborn could be both preterm *and* delivered via Cesarean section, although the model in our analysis separates the contributions of these two factors.

Third, our results also highlighted the perils of early-life uses of anti-infective. A general trend of decreased microbial diversity due to antibiotics use has been observed in children. The resulting dysbiosis is now known to be associated with a wide range of immune system-related diseases. Finally, though we observe significant sex differences in the expected count for a few infections and immune disorders, not all of them agreed with previous studies about adult, sex-specific disease rates.^36^ Because adult sex differences are linked to post-puberty hormonal levels, there is no real discrepancy between our results and previous studies, due to the difference in cohorts.^37,38^

This study examines the association between immune-system-related diseases and perinatal and postnatal health factors. Our results suggest that some potential risk factors are, indeed, associated with significant changes in neonatal diseases and thus deserve additional attention from clinicians and society at large.

## Data Availability

The IBM MarketScan data is available by to licensed users. See https://www.ibm.com/products/marketscan-research-databases

## Acknowledgments

The authors are grateful to E. Gannon and M. Rzhetsky for comments on earlier versions of the manuscript. This work was funded by the DARPA Big Mechanism program under ARO contract W911NF1410333, by National Institutes of Health grants R01HL122712, 1P50MH094267, and U01HL108634-01, and by a gift from Liz and Kent Dauten. The funders had no role in study design, data collection and analysis, decision to publish, or preparation of the manuscript. The funders had no role in study design, data collection and analysis, decision to publish, or preparation of the man uscript.

## Notes

### Competing Interest Statement

The authors have declared no competing interest.

### Author Declarations

The University of Chicago Institutional Review Boards determined that the study is IRB exempt, given that patient data in both countries were preexisting and de-identified.

### Summary of Updates

Main text was updated to clarify the reporting of results.

