## Supplementary information for "Peri- and Post-natal Risk Factors Associated with Health of Newborns"

January 9, 2023

### 1 Methods

#### 1.1 Data Source and Preprocessing

In this study, we used IBM MarketScan data from 2003–2018 [2], which contained daily-level information on diagnosis (DX), prescription (RX), procedure (PX), laboratory tests, and household information for more than 150 million unique subjects from the United States. Using the latter we computed mother–newborn links similar to previous studies (e.g. [4, 3]). Specifically, we attempted to match each newborn subject, defined as anyone having at least one delivery-related DX code under the International Classification of Diseases (ICD, versions 9 and 10) [10] at age 0, with a female within the same household also having at least one delivery-related DX code within 3 days of the day on which the first newborn delivery-related DX code was registered, which was treated in subsequent analyses as the birthday of the said newborn. We required newborns to have been followed continuously from birth for at least 1 year, allowing up to 1 month of lapse between birth and first day of enrolment, since in many cases the enrolment period started only on the first

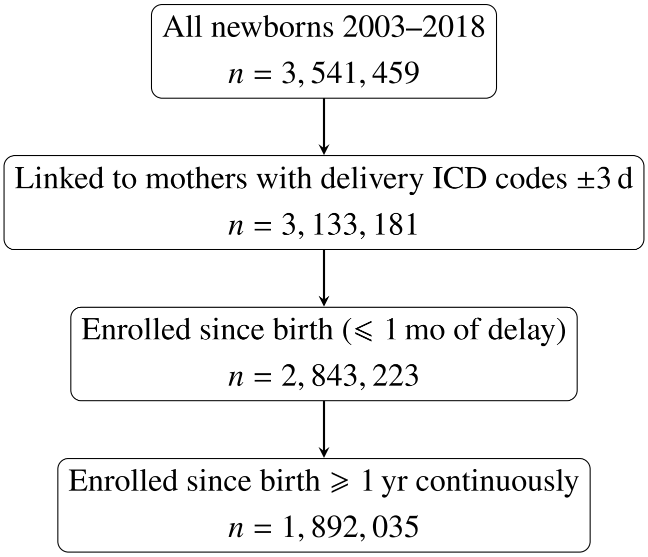


Figure S1: Flowchart for inclusion/exclusion of subjects

day of the month following the month of birth. This inclusion/exclusion procedure is succinctly represented in a flowchart (Figure S1).

In this study, we investigated two classes of diseases that involve the immune system: infections and immune disorders. We split infections into sub-phenotypes: bacterial, viral, miscellaneous (fungi, protozoans, worms, and infestations) and sequaelae. The ICD codes used for each phenotype are listed in a separate text file. We fitted Bayesian negative binomial (NB) regression models with newborn diseases counts as outcome, grouping together observations with the same values in the predictors and summing up the correponding values in the outcomes. Possible predictors include the same phenotypes (DXs) for the linked mothers, antiinfective RXs for mothers up to birth and for newborns during the first year of life, plus additional peri- and neo-natal conditions (Table S1). Specifically, all models include predictors $C$ and $P$ and all combinations from the set $\{W, A, R\}$. The predictor DXs and RXs were modelled as binary variables, indicating the presence/absence of the corresponding events. Finally, for each combination we fitted both hurdle (H) and non-hurdle versions of the models.

| Abbreviation | code | description |
|---|---|---|
| $C$ | `csec` | Caesarean section |
| $P$ | `prem` | Preterm birth: $<$ 37 wk of gestation |
| $W$ | `weight` | Birthweight: low ($\leqslant$ 2500 g), high ($\geqslant$ 4500 g) |
| $A$ | `age` | Mother's age at birth: teenage (13–18), advanced (35–45) |
| $R$ | `RX` | Antiinfective prescriptions (antibacterials, antimycotics, antiparasitics) |
| $H$ | `hurd` | Hurdle |

Table S1: Abbreviations for additional model terms.

#### 1.2 Regression Analysis

The regression equation is given by [9, §§4.2.1 & 4.3, pp. 131 & 134]

$$y_i \sim \mathrm{NB}\left(r, \exp\left(\mathbf{x}^\top \boldsymbol{\beta}\right)\right) \iff \\ \mathbb{P}_{\mathrm{NB}}(y_i|\mathbf{x}, \boldsymbol{\beta}, r) = \frac{\Gamma(y_i + r)}{\Gamma(y_i - 1)\Gamma(r)} \left[\frac{\exp\left(\mathbf{x}^\top \boldsymbol{\beta}\right)}{\exp\left(\mathbf{x}^\top \boldsymbol{\beta}\right) + r}\right]^{y_i} \left[\frac{r}{\exp\left(\mathbf{x}^\top \boldsymbol{\beta}\right) + r}\right]^{r}. \tag{1}$$

In count data regression, a regression coefficient (elements of $\boldsymbol{\beta}$) represents the relative effect on the expected counts of the outcome variable due to 1 unit increase of the corresponding predictor, or, in the case of binary predictors, the presence over the absence of the corresponding phenotype. Formally, the relative change in expected count, $\mathbb{E}\left(y|\cdot\right)$ (relative count change, $\Delta$RC), due to such a change in $\mathbf{x}_j$ w.r.t. the baseline can be written as follows [9, §3.1.4, pp. 70–71]

$$\begin{aligned} \Delta\mathrm{RC}_j &= \frac{\mathbb{E}\left(y|\,\mathbf{x}^{(j)\top}\boldsymbol{\beta}\right) - \mathbb{E}\left(y|\,\mathbf{x}^\top\boldsymbol{\beta}\right)}{\mathbb{E}\left(y|\,\mathbf{x}^\top\boldsymbol{\beta}\right)} \\ &= \frac{\exp\left(\mathbf{x}^\top\boldsymbol{\beta} + \boldsymbol{\beta}_j\right) - \exp\left(\mathbf{x}^\top\boldsymbol{\beta}\right)}{\exp\left(\mathbf{x}^\top\boldsymbol{\beta}\right)} \\ &= \exp\left(\boldsymbol{\beta}_j\right) - 1, \end{aligned} \tag{2}$$

where $\mathbf{x}^{(j)}$ is the same as $\mathbf{x}$ except that its $j^{\text{th}}$ entry is increased by 1, and $\boldsymbol{\beta}_j$ the $j^{\text{th}}$ entry of $\boldsymbol{\beta}_j$.

As mentioned before, in addition to the vanilla NB models we also fitted hurdle NB models, with likelihood given by

$$\mathbb{P}_{\mathrm{Hurdle-NB}}(y_i|\mathbf{x}, \boldsymbol{\beta}, r) = \begin{cases} \pi & y_i = 0 \\ (1 - \pi)\,\dfrac{\mathbb{P}_{\mathrm{NB}}(y_i|\mathbf{x}, \boldsymbol{\beta}, r)}{1 - \mathbb{P}_{\mathrm{NB}}(0|\mathbf{x}, \boldsymbol{\beta}, r)} & y_i > 0, \end{cases} \tag{3}$$

where $\pi \in [0, 1]$ represent the proportion of the data points $\{y_i\}$ equal to 0. As such, the hurdle model can be used to both inflate and deflate zero outcomes. For the prior on $\pi$ we used $\mathrm{Unif}(0, 1) \equiv \mathrm{Beta}(1, 1)$.

#### 1.3 Statistical Modelling

##### 1.3.1 Over-dispersion

Table S2 (see also Figure S7) shows that for each of the outcome phenotypes the variance was much larger than the mean, suggesting the presence of strong over-dispersion with respect to the Poisson distribution.

##### 1.3.2 MCMC Convergence

Following [8], for each variable and for the log-posterior we checked if $\hat{R} < 1.01$ and if rank-normalized bulk- and tail-estimated sample size (ESS) are greater than 400. This would ensure

| Phenotype | Median | Mean | Variance | Variance/Mean |
|---|---|---|---|---|
| Infections | | | | |
| Bacterial | 0 | 0.237 | 1.61 | 6.8 |
| Viral | 0 | 0.522 | 1.7 | 3.25 |
| Miscellaneous | 0 | 0.238 | 0.911 | 3.83 |
| Sequelae | 0 | 0.802 | 2.49 | 3.1 |
| Immune disorder | 2 | 5.49 | 181 | 32.9 |

Table S2: Summary statistics of counts of the outcomes

good inter- *and* intra-chain mixing. In addition, we performed graphical checks by inspecting the trace and pairs plot as well as the rank plot, which shows if of the ranks of the posterior draws are similar across the chains. An equivalent way to check this is to see if the empirical cumulative distribution function (ECDF) of the ranks represents a sample from a uniform distribution [5].

###### 1.3.3 Model Comparison

We first approximated leave-one-out (LOO) cross-validation via Pareto smoothed importance sampling (PSIS)[6], which computes the expected log pointwise predictive density for new data points, given by

$$\mathrm{ELPD}_{\mathrm{loo}} = \sum_{i=0}^{N} \log \underbrace{\int p(y_i|\boldsymbol{\theta})p(\boldsymbol{\theta}|y_{-i})\,d\boldsymbol{\theta}}_{p(y_i|y_{i-1})}, \tag{4}$$

by drawing samples of $\boldsymbol{\theta}$ from the posterior using importance sampling with importance ratios

$$r_i^{(s)} = \frac{1}{p\left(y_i|\boldsymbol{\theta}^{(s)}\right)}, \tag{5}$$

smoothing data points whose $r^{(s)}$ fall in the top 20% with the quantile function of the generalized Pareto distribution, truncating the weights to ensure infinite variances, and finally evaluating the integrand in (4)

$$p(y_i|y_{i-1}) \approx \frac{S}{\sum_{s=1}^{S} r_i^{(s)}} \tag{6}$$

However, if the shape parameter $\hat{k}$ of the fitted generalized Pareto distribution is greater than 0.7, the corresponding data point is considered "problematic", and if the number of problematic data points is large, $K$-fold cross-validation (CV) would be more robust than PSIS-LOO [7, 6]. Since we observed more than 10 problematic data points, we shall report results from 10-fold CV instead. The dots in the plots represent the differences in ELPD whereas the line segments represent the 95% credible intervals.

| model | elpd_diff | se_diff | elpd_kfold | se_elpd_kfold | p_kfold | se_p_kfold |
|---|---|---|---|---|---|---|
| C + P + W + A + R / H | $0.000 \times 10^0$ | $0.000 \times 10^0$ | $-1.917 \times 10^5$ | $6.680 \times 10^2$ | $-1.836 \times 10^5$ | $5.453 \times 10^3$ |
| C + P + A + R / H | $-2.373 \times 10^2$ | $3.683 \times 10^1$ | $-1.920 \times 10^5$ | $6.710 \times 10^2$ | $-1.761 \times 10^5$ | $5.180 \times 10^3$ |
| C + P + W + R / H | $-2.490 \times 10^2$ | $3.863 \times 10^1$ | $-1.920 \times 10^5$ | $6.723 \times 10^2$ | $-1.834 \times 10^5$ | $5.447 \times 10^3$ |
| C + P + W + A + R | $-3.860 \times 10^2$ | $1.150 \times 10^2$ | $-1.921 \times 10^5$ | $6.957 \times 10^2$ | $-2.707 \times 10^5$ | $6.588 \times 10^3$ |
| C + P + R / H | $-4.652 \times 10^2$ | $5.361 \times 10^1$ | $-1.922 \times 10^5$ | $6.744 \times 10^2$ | $-1.760 \times 10^5$ | $5.184 \times 10^3$ |
| C + P + W + A / H | $-7.571 \times 10^2$ | $7.905 \times 10^1$ | $-1.925 \times 10^5$ | $6.752 \times 10^2$ | $-1.757 \times 10^5$ | $5.169 \times 10^3$ |
| C + P + A + R | $-8.355 \times 10^2$ | $1.229 \times 10^2$ | $-1.926 \times 10^5$ | $6.990 \times 10^2$ | $-2.676 \times 10^5$ | $6.484 \times 10^3$ |
| C + P + W + R | $-8.362 \times 10^2$ | $1.278 \times 10^2$ | $-1.926 \times 10^5$ | $7.007 \times 10^2$ | $-2.710 \times 10^5$ | $6.623 \times 10^3$ |
| C + P + W / H | $-9.685 \times 10^2$ | $9.482 \times 10^1$ | $-1.927 \times 10^5$ | $6.789 \times 10^2$ | $-1.756 \times 10^5$ | $5.172 \times 10^3$ |
| C + P + A / H | $-1.000 \times 10^3$ | $8.897 \times 10^1$ | $-1.927 \times 10^5$ | $6.779 \times 10^2$ | $-1.685 \times 10^5$ | $4.903 \times 10^3$ |
| C + P / H | $-1.250 \times 10^3$ | $1.062 \times 10^2$ | $-1.930 \times 10^5$ | $6.838 \times 10^2$ | $-1.684 \times 10^5$ | $4.903 \times 10^3$ |
| C + P + R | $-1.309 \times 10^3$ | $1.382 \times 10^2$ | $-1.930 \times 10^5$ | $7.055 \times 10^2$ | $-2.679 \times 10^5$ | $6.524 \times 10^3$ |
| C + P + W + A | $-1.525 \times 10^3$ | $1.420 \times 10^2$ | $-1.933 \times 10^5$ | $7.039 \times 10^2$ | $-2.658 \times 10^5$ | $6.412 \times 10^3$ |
| C + P + W | $-1.961 \times 10^3$ | $1.580 \times 10^2$ | $-1.937 \times 10^5$ | $7.111 \times 10^2$ | $-2.662 \times 10^5$ | $6.450 \times 10^3$ |
| C + P + A | $-1.966 \times 10^3$ | $1.516 \times 10^2$ | $-1.937 \times 10^5$ | $7.090 \times 10^2$ | $-2.630 \times 10^5$ | $6.316 \times 10^3$ |
| C + P | $-2.418 \times 10^3$ | $1.685 \times 10^2$ | $-1.941 \times 10^5$ | $7.171 \times 10^2$ | $-2.634 \times 10^5$ | $6.358 \times 10^3$ |

Table S3: Bacterial

Figure S2: Bacterial

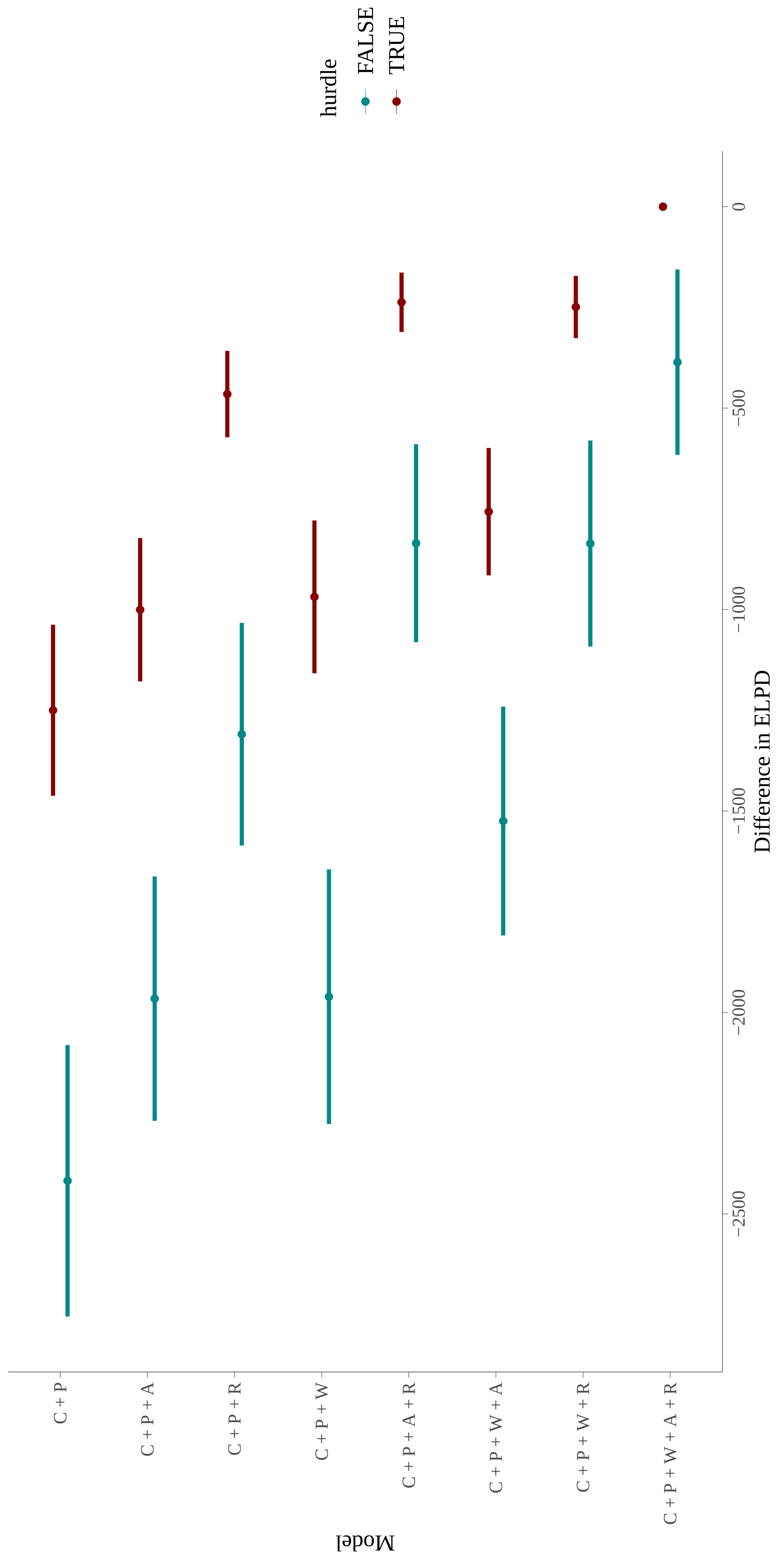

| model | elpd_diff | se_diff | elpd_kfold | se_elpd_kfold | p_kfold | se_p_kfold |
|---|---|---|---|---|---|---|
| C + P + W + A + R / H | $0.000 \times 10^{0}$ | $0.000 \times 10^{0}$ | $-7.131 \times 10^{5}$ | $8.716 \times 10^{2}$ | $-1.681 \times 10^{6}$ | $3.366 \times 10^{4}$ |
| C + P + W + R / H | $-5.472 \times 10^{2}$ | $5.107 \times 10^{1}$ | $-7.136 \times 10^{5}$ | $8.766 \times 10^{2}$ | $-1.687 \times 10^{6}$ | $3.394 \times 10^{4}$ |
| C + P + A + R / H | $-7.551 \times 10^{2}$ | $4.610 \times 10^{1}$ | $-7.138 \times 10^{5}$ | $8.770 \times 10^{2}$ | $-1.648 \times 10^{6}$ | $3.277 \times 10^{4}$ |
| C + P + W + A / H | $-1.114 \times 10^{3}$ | $6.637 \times 10^{1}$ | $-7.142 \times 10^{5}$ | $8.762 \times 10^{2}$ | $-1.583 \times 10^{6}$ | $3.046 \times 10^{4}$ |
| C + P + R / H | $-1.311 \times 10^{3}$ | $6.919 \times 10^{1}$ | $-7.144 \times 10^{5}$ | $8.827 \times 10^{2}$ | $-1.654 \times 10^{6}$ | $3.301 \times 10^{4}$ |
| C + P + W / H | $-1.654 \times 10^{3}$ | $8.363 \times 10^{1}$ | $-7.147 \times 10^{5}$ | $8.821 \times 10^{2}$ | $-1.589 \times 10^{6}$ | $3.074 \times 10^{4}$ |
| C + P + A / H | $-1.892 \times 10^{3}$ | $8.068 \times 10^{1}$ | $-7.149 \times 10^{5}$ | $8.828 \times 10^{2}$ | $-1.557 \times 10^{6}$ | $2.978 \times 10^{4}$ |
| C + P / H | $-2.444 \times 10^{3}$ | $9.666 \times 10^{1}$ | $-7.155 \times 10^{5}$ | $8.892 \times 10^{2}$ | $-1.563 \times 10^{6}$ | $3.006 \times 10^{4}$ |
| C + P + W + A + R | $-1.176 \times 10^{4}$ | $3.184 \times 10^{2}$ | $-7.248 \times 10^{5}$ | $1.035 \times 10^{3}$ | $-1.875 \times 10^{6}$ | $3.680 \times 10^{4}$ |
| C + P + W + R | $-1.281 \times 10^{4}$ | $3.463 \times 10^{2}$ | $-7.259 \times 10^{5}$ | $1.052 \times 10^{3}$ | $-1.881 \times 10^{6}$ | $3.710 \times 10^{4}$ |
| C + P + A + R | $-1.308 \times 10^{4}$ | $3.461 \times 10^{2}$ | $-7.261 \times 10^{5}$ | $1.054 \times 10^{3}$ | $-1.844 \times 10^{6}$ | $3.591 \times 10^{4}$ |
| C + P + W + A | $-1.380 \times 10^{4}$ | $3.497 \times 10^{2}$ | $-7.269 \times 10^{5}$ | $1.054 \times 10^{3}$ | $-1.775 \times 10^{6}$ | $3.339 \times 10^{4}$ |
| C + P + R | $-1.417 \times 10^{4}$ | $3.754 \times 10^{2}$ | $-7.272 \times 10^{5}$ | $1.072 \times 10^{3}$ | $-1.852 \times 10^{6}$ | $3.627 \times 10^{4}$ |
| C + P + W | $-1.484 \times 10^{4}$ | $3.799 \times 10^{2}$ | $-7.279 \times 10^{5}$ | $1.073 \times 10^{3}$ | $-1.782 \times 10^{6}$ | $3.370 \times 10^{4}$ |
| C + P + A | $-1.512 \times 10^{4}$ | $3.794 \times 10^{2}$ | $-7.282 \times 10^{5}$ | $1.075 \times 10^{3}$ | $-1.750 \times 10^{6}$ | $3.270 \times 10^{4}$ |
| C + P | $-1.618 \times 10^{4}$ | $4.100 \times 10^{2}$ | $-7.292 \times 10^{5}$ | $1.095 \times 10^{3}$ | $-1.758 \times 10^{6}$ | $3.302 \times 10^{4}$ |

Table S4: Immune

Figure S3: Immune

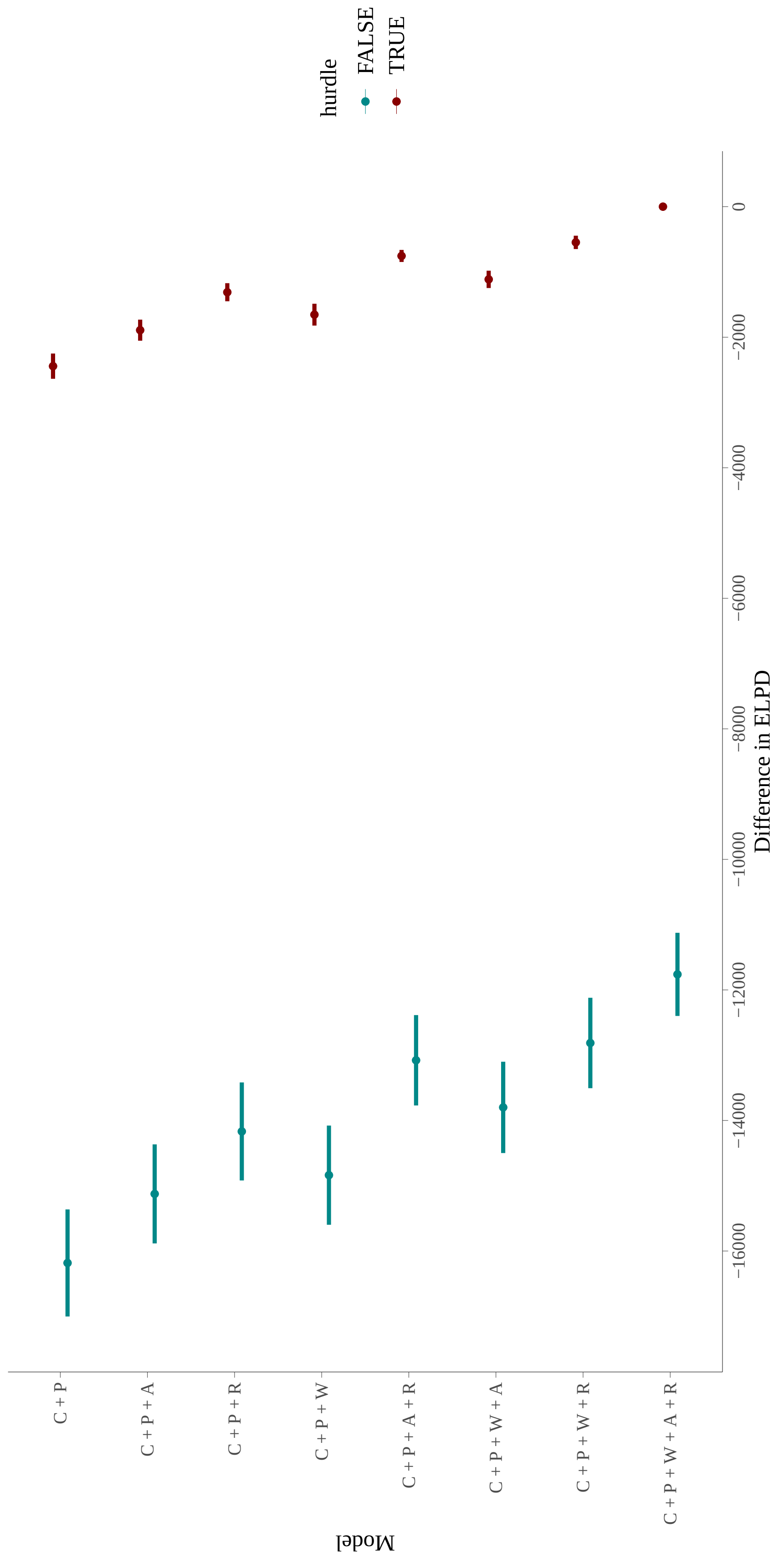

| model | elpd_diff | se_diff | elpd_kfold | se_elpd_kfold | p_kfold | se_p_kfold |
|---|---|---|---|---|---|---|
| C + P + W + A + R | $0.000 \times 10^{0}$ | $0.000 \times 10^{0}$ | $-1.931 \times 10^{5}$ | $6.828 \times 10^{2}$ | $-2.235 \times 10^{5}$ | $4.926 \times 10^{3}$ |
| C + P + W + A + R / H | $-3.982 \times 10^{1}$ | $1.199 \times 10^{2}$ | $-1.931 \times 10^{5}$ | $6.499 \times 10^{2}$ | $-1.406 \times 10^{5}$ | $3.967 \times 10^{3}$ |
| C + P + W + R / H | $-2.527 \times 10^{2}$ | $1.195 \times 10^{2}$ | $-1.933 \times 10^{5}$ | $6.515 \times 10^{2}$ | $-1.402 \times 10^{5}$ | $3.965 \times 10^{3}$ |
| C + P + A + R / H | $-3.187 \times 10^{2}$ | $1.269 \times 10^{2}$ | $-1.934 \times 10^{5}$ | $6.472 \times 10^{2}$ | $-1.383 \times 10^{5}$ | $3.900 \times 10^{3}$ |
| C + P + W + R | $-4.385 \times 10^{2}$ | $4.699 \times 10^{1}$ | $-1.935 \times 10^{5}$ | $6.856 \times 10^{2}$ | $-2.236 \times 10^{5}$ | $4.952 \times 10^{3}$ |
| C + P + R / H | $-5.548 \times 10^{2}$ | $1.303 \times 10^{2}$ | $-1.936 \times 10^{5}$ | $6.512 \times 10^{2}$ | $-1.378 \times 10^{5}$ | $3.882 \times 10^{3}$ |
| C + P + A + R | $-6.359 \times 10^{2}$ | $5.219 \times 10^{1}$ | $-1.937 \times 10^{5}$ | $6.833 \times 10^{2}$ | $-2.224 \times 10^{5}$ | $4.907 \times 10^{3}$ |
| C + P + W + A / H | $-9.414 \times 10^{2}$ | $1.183 \times 10^{2}$ | $-1.940 \times 10^{5}$ | $6.576 \times 10^{2}$ | $-1.367 \times 10^{5}$ | $3.858 \times 10^{3}$ |
| C + P + R | $-1.127 \times 10^{3}$ | $8.176 \times 10^{1}$ | $-1.942 \times 10^{5}$ | $6.901 \times 10^{2}$ | $-2.225 \times 10^{5}$ | $4.929 \times 10^{3}$ |
| C + P + W / H | $-1.148 \times 10^{3}$ | $1.236 \times 10^{2}$ | $-1.942 \times 10^{5}$ | $6.611 \times 10^{2}$ | $-1.364 \times 10^{5}$ | $3.857 \times 10^{3}$ |
| C + P + W + A | $-1.180 \times 10^{3}$ | $7.187 \times 10^{1}$ | $-1.943 \times 10^{5}$ | $6.954 \times 10^{2}$ | $-2.198 \times 10^{5}$ | $4.832 \times 10^{3}$ |
| C + P + A / H | $-1.240 \times 10^{3}$ | $1.280 \times 10^{2}$ | $-1.943 \times 10^{5}$ | $6.609 \times 10^{2}$ | $-1.345 \times 10^{5}$ | $3.796 \times 10^{3}$ |
| C + P / H | $-1.455 \times 10^{3}$ | $1.354 \times 10^{2}$ | $-1.945 \times 10^{5}$ | $6.657 \times 10^{2}$ | $-1.341 \times 10^{5}$ | $3.776 \times 10^{3}$ |
| C + P + W | $-1.592 \times 10^{3}$ | $9.193 \times 10^{1}$ | $-1.947 \times 10^{5}$ | $6.998 \times 10^{2}$ | $-2.199 \times 10^{5}$ | $4.856 \times 10^{3}$ |
| C + P + A | $-1.782 \times 10^{3}$ | $9.506 \times 10^{1}$ | $-1.949 \times 10^{5}$ | $6.979 \times 10^{2}$ | $-2.188 \times 10^{5}$ | $4.811 \times 10^{3}$ |
| C + P | $-2.239 \times 10^{3}$ | $1.169 \times 10^{2}$ | $-1.953 \times 10^{5}$ | $7.057 \times 10^{2}$ | $-2.189 \times 10^{5}$ | $4.835 \times 10^{3}$ |

Table S5: Miscellaneous

| model | elpd_diff | se_diff | elpd_kfold | se_elpd_kfold | p_kfold | se_p_kfold |
|---|---|---|---|---|---|---|
| C + C + P + P + W + W + A + A + R + R | $0.000 \times 10^{0}$ | $0.000 \times 10^{0}$ | $-1.931 \times 10^{5}$ | $6.828 \times 10^{2}$ | $-2.235 \times 10^{5}$ | $4.926 \times 10^{3}$ |
| C + C + P + P + W + W + A + A + R + R / H | $-3.982 \times 10^{1}$ | $1.199 \times 10^{2}$ | $-1.931 \times 10^{5}$ | $6.499 \times 10^{2}$ | $-1.406 \times 10^{5}$ | $3.967 \times 10^{3}$ |

Table S6: Miscellaneous

Figure S4: Miscellaneous

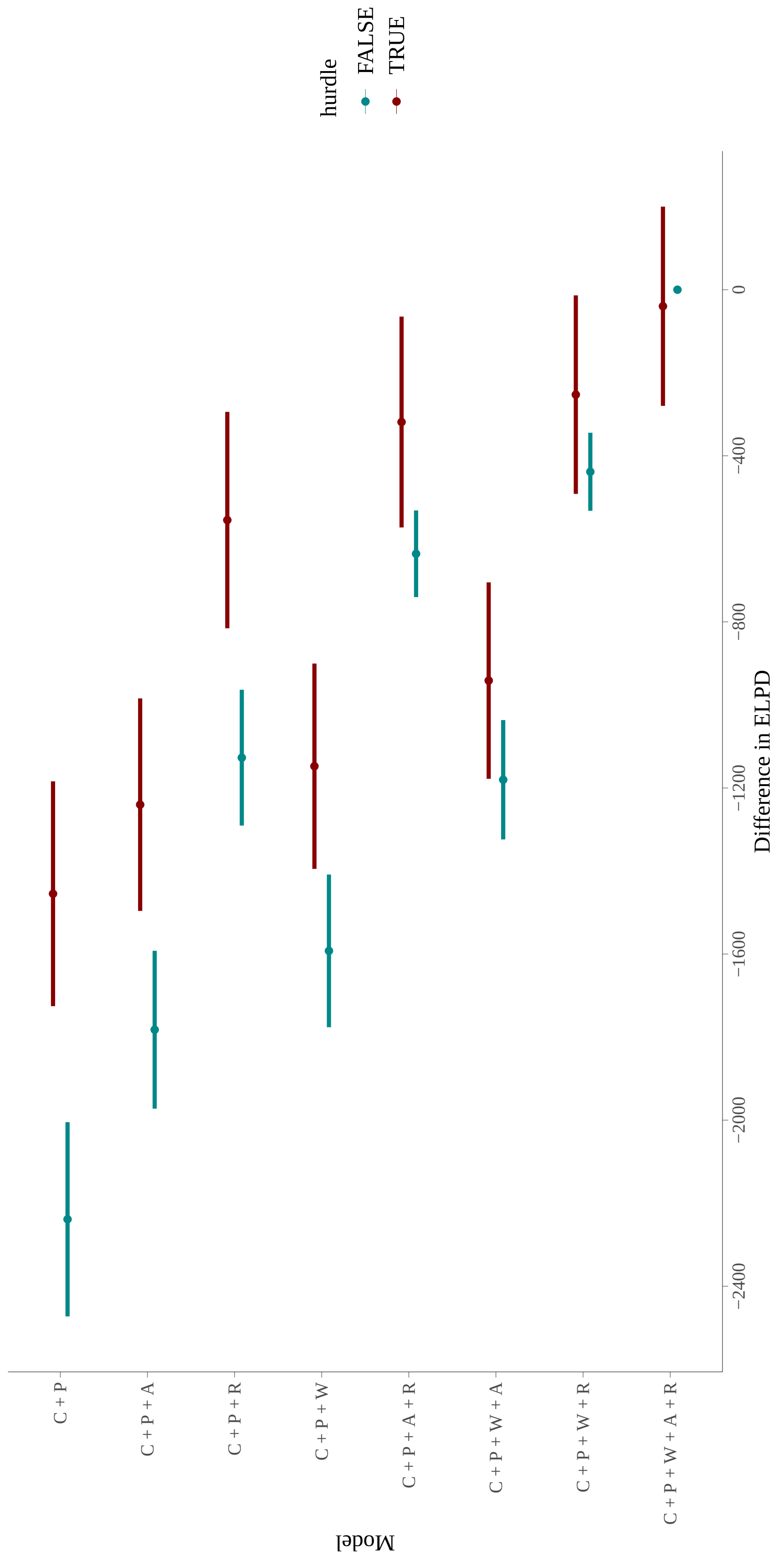

| model | elpd_diff | se_diff | elpd_kfold | se_elpd_kfold | p_kfold | se_p_kfold |
|---|---|---|---|---|---|---|
| C + P + W + A + R / H | $0.000 \times 10^{0}$ | $0.000 \times 10^{0}$ | $-3.664 \times 10^{5}$ | $7.472 \times 10^{2}$ | $-7.747 \times 10^{5}$ | $1.959 \times 10^{4}$ |
| C + P + W + R / H | $-3.540 \times 10^{2}$ | $4.124 \times 10^{1}$ | $-3.668 \times 10^{5}$ | $7.541 \times 10^{2}$ | $-7.711 \times 10^{5}$ | $1.938 \times 10^{4}$ |
| C + P + A + R / H | $-7.159 \times 10^{2}$ | $4.724 \times 10^{1}$ | $-3.671 \times 10^{5}$ | $7.558 \times 10^{2}$ | $-7.607 \times 10^{5}$ | $1.914 \times 10^{4}$ |
| C + P + R / H | $-1.145 \times 10^{3}$ | $7.026 \times 10^{1}$ | $-3.676 \times 10^{5}$ | $7.657 \times 10^{2}$ | $-7.581 \times 10^{5}$ | $1.903 \times 10^{4}$ |
| C + P + W + A / H | $-2.012 \times 10^{3}$ | $9.265 \times 10^{1}$ | $-3.684 \times 10^{5}$ | $7.682 \times 10^{2}$ | $-7.602 \times 10^{5}$ | $1.918 \times 10^{4}$ |
| C + P + A / H | $-2.826 \times 10^{3}$ | $1.140 \times 10^{2}$ | $-3.692 \times 10^{5}$ | $7.800 \times 10^{2}$ | $-7.471 \times 10^{5}$ | $1.877 \times 10^{4}$ |
| C + P / H | $-3.213 \times 10^{3}$ | $1.313 \times 10^{2}$ | $-3.696 \times 10^{5}$ | $7.896 \times 10^{2}$ | $-7.451 \times 10^{5}$ | $1.865 \times 10^{4}$ |
| C + P + W + A + R | $-8.024 \times 10^{3}$ | $2.441 \times 10^{2}$ | $-3.744 \times 10^{5}$ | $8.739 \times 10^{2}$ | $-8.482 \times 10^{5}$ | $1.949 \times 10^{4}$ |
| C + P + W + R | $-8.694 \times 10^{3}$ | $2.658 \times 10^{2}$ | $-3.751 \times 10^{5}$ | $8.875 \times 10^{2}$ | $-8.467 \times 10^{5}$ | $1.940 \times 10^{4}$ |
| C + P + A + R | $-9.145 \times 10^{3}$ | $2.673 \times 10^{2}$ | $-3.756 \times 10^{5}$ | $8.900 \times 10^{2}$ | $-8.383 \times 10^{5}$ | $1.917 \times 10^{4}$ |
| C + P + R | $-9.815 \times 10^{3}$ | $2.900 \times 10^{2}$ | $-3.762 \times 10^{5}$ | $9.046 \times 10^{2}$ | $-8.370 \times 10^{5}$ | $1.910 \times 10^{4}$ |
| C + P + W + A | $-1.103 \times 10^{4}$ | $3.014 \times 10^{2}$ | $-3.774 \times 10^{5}$ | $9.114 \times 10^{2}$ | $-8.367 \times 10^{5}$ | $1.913 \times 10^{4}$ |
| C + P + W | $-1.161 \times 10^{4}$ | $3.210 \times 10^{2}$ | $-3.780 \times 10^{5}$ | $9.245 \times 10^{2}$ | $-8.363 \times 10^{5}$ | $1.913 \times 10^{4}$ |
| C + P + A | $-1.211 \times 10^{4}$ | $3.250 \times 10^{2}$ | $-3.785 \times 10^{5}$ | $9.293 \times 10^{2}$ | $-8.279 \times 10^{5}$ | $1.887 \times 10^{4}$ |
| C + P | $-1.271 \times 10^{4}$ | $3.466 \times 10^{2}$ | $-3.791 \times 10^{5}$ | $9.439 \times 10^{2}$ | $-8.273 \times 10^{5}$ | $1.883 \times 10^{4}$ |
| C + P + W / H | – | – | – | – | – | – |

Table S7: Sequelae

Figure S5: Sequelae

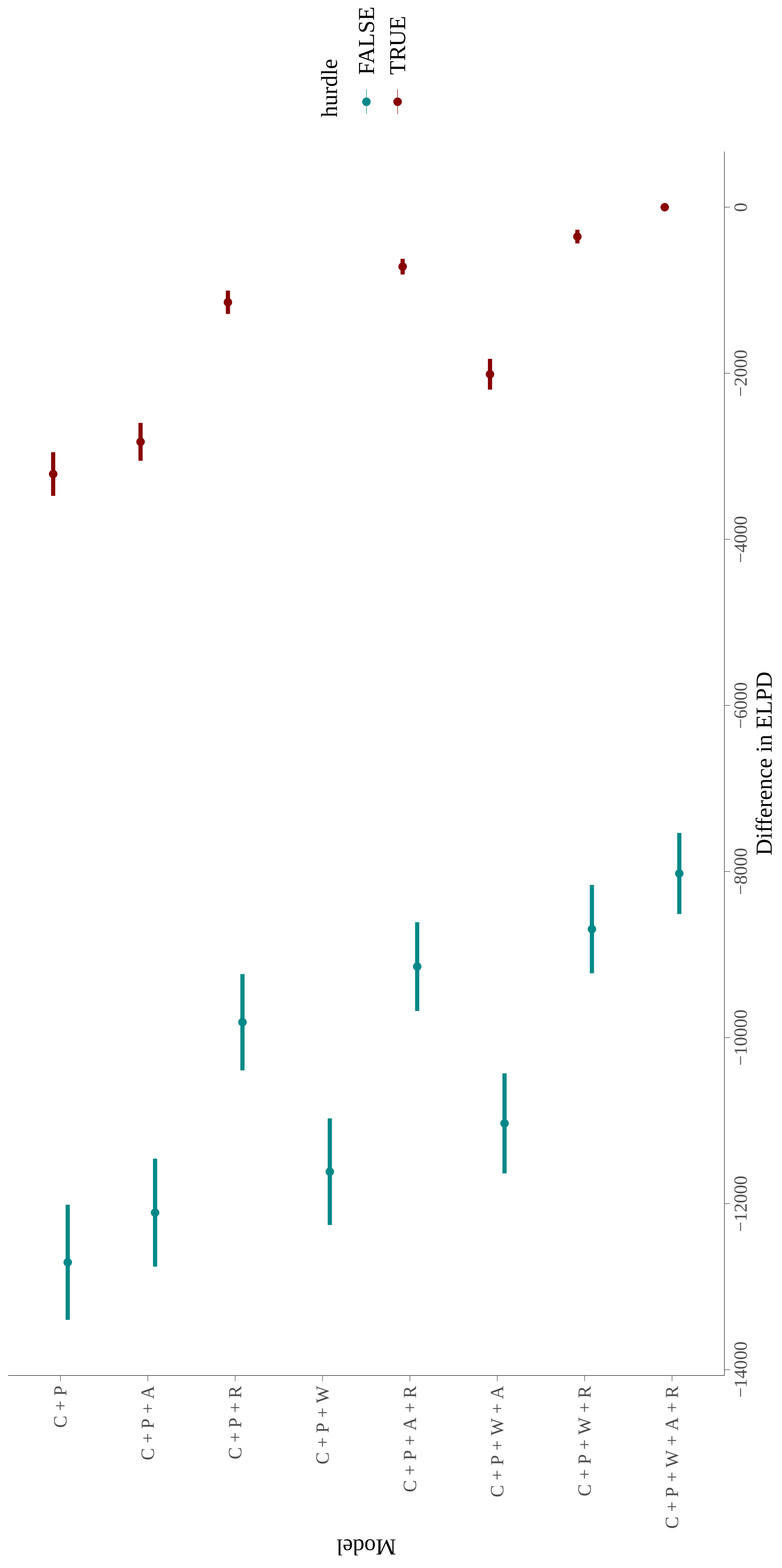

| model | elpd_diff | se_diff | elpd_kfold | se_elpd_kfold | p_kfold | se_p_kfold |
|---|---|---|---|---|---|---|
| C + P + W + A + R / H | $0.000 \times 10^{0}$ | $0.000 \times 10^{0}$ | $-3.165 \times 10^{5}$ | $7.327 \times 10^{2}$ | $-5.872 \times 10^{5}$ | $1.411 \times 10^{4}$ |
| C + P + W + R / H | $-3.068 \times 10^{2}$ | $3.836 \times 10^{1}$ | $-3.169 \times 10^{5}$ | $7.379 \times 10^{2}$ | $-5.859 \times 10^{5}$ | $1.403 \times 10^{4}$ |
| C + P + A + R / H | $-7.096 \times 10^{2}$ | $4.998 \times 10^{1}$ | $-3.173 \times 10^{5}$ | $7.390 \times 10^{2}$ | $-5.827 \times 10^{5}$ | $1.397 \times 10^{4}$ |
| C + P + R / H | $-1.027 \times 10^{3}$ | $6.580 \times 10^{1}$ | $-3.176 \times 10^{5}$ | $7.443 \times 10^{2}$ | $-5.816 \times 10^{5}$ | $1.391 \times 10^{4}$ |
| C + P + W + A / H | $-1.269 \times 10^{3}$ | $7.531 \times 10^{1}$ | $-3.178 \times 10^{5}$ | $7.433 \times 10^{2}$ | $-5.698 \times 10^{5}$ | $1.350 \times 10^{4}$ |
| C + P + W / H | $-1.567 \times 10^{3}$ | $8.725 \times 10^{1}$ | $-3.181 \times 10^{5}$ | $7.486 \times 10^{2}$ | $-5.691 \times 10^{5}$ | $1.346 \times 10^{4}$ |
| C + P + A / H | $-2.023 \times 10^{3}$ | $9.450 \times 10^{1}$ | $-3.186 \times 10^{5}$ | $7.514 \times 10^{2}$ | $-5.657 \times 10^{5}$ | $1.337 \times 10^{4}$ |
| C + P / H | $-2.341 \times 10^{3}$ | $1.084 \times 10^{2}$ | $-3.189 \times 10^{5}$ | $7.578 \times 10^{2}$ | $-5.653 \times 10^{5}$ | $1.336 \times 10^{4}$ |
| C + P + W + A + R | $-4.160 \times 10^{3}$ | $1.820 \times 10^{2}$ | $-3.207 \times 10^{5}$ | $8.083 \times 10^{2}$ | $-6.714 \times 10^{5}$ | $1.436 \times 10^{4}$ |
| C + P + W + R | $-4.799 \times 10^{3}$ | $1.991 \times 10^{2}$ | $-3.213 \times 10^{5}$ | $8.179 \times 10^{2}$ | $-6.717 \times 10^{5}$ | $1.439 \times 10^{4}$ |
| C + P + A + R | $-5.352 \times 10^{3}$ | $2.014 \times 10^{2}$ | $-3.219 \times 10^{5}$ | $8.202 \times 10^{2}$ | $-6.689 \times 10^{5}$ | $1.430 \times 10^{4}$ |
| C + P + R | $-6.002 \times 10^{3}$ | $2.194 \times 10^{2}$ | $-3.226 \times 10^{5}$ | $8.306 \times 10^{2}$ | $-6.689 \times 10^{5}$ | $1.429 \times 10^{4}$ |
| C + P + W + A | $-6.033 \times 10^{3}$ | $2.150 \times 10^{2}$ | $-3.226 \times 10^{5}$ | $8.266 \times 10^{2}$ | $-6.587 \times 10^{5}$ | $1.390 \times 10^{4}$ |
| C + P + W | $-6.625 \times 10^{3}$ | $2.320 \times 10^{2}$ | $-3.232 \times 10^{5}$ | $8.368 \times 10^{2}$ | $-6.590 \times 10^{5}$ | $1.392 \times 10^{4}$ |
| C + P + A | $-7.188 \times 10^{3}$ | $2.346 \times 10^{2}$ | $-3.237 \times 10^{5}$ | $8.397 \times 10^{2}$ | $-6.566 \times 10^{5}$ | $1.385 \times 10^{4}$ |
| C + P | $-7.801 \times 10^{3}$ | $2.533 \times 10^{2}$ | $-3.244 \times 10^{5}$ | $8.512 \times 10^{2}$ | $-6.569 \times 10^{5}$ | $1.388 \times 10^{4}$ |

Table S8: Viral

Figure S6: Viral

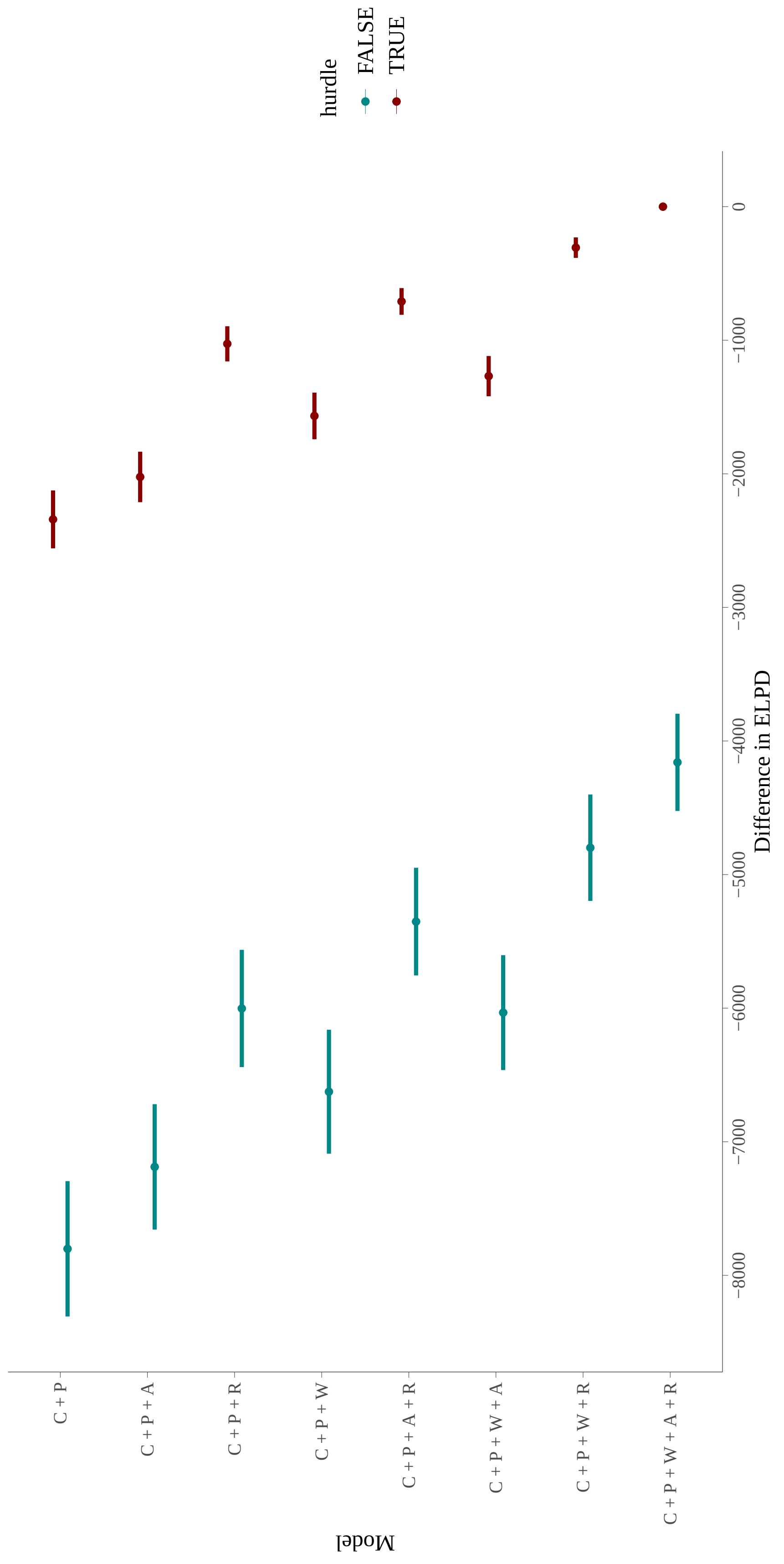

#### 1.4 Proofs

We show below that the sum of i.i.d. NB random variables is an NB random variable with the same mean parameter and with shape parameter added together, which justifies our use of grouped data. Another way to do so is to note that since the NB is an exponential family distribution (with fixed but unknown $r$), the sufficient statistics for the set of parameters $\left\{\boldsymbol{\beta}_j\right\}$ when using the canonical link function are the entries of $\mathbf{x}^\top \mathbf{y}$ [1, §4.2, pp. 123–124], which is the same for grouped and ungrouped data.

The characteristic function of $Y \sim \mathrm{NB}\,(r, p)$ is given by

$$\begin{aligned}
\phi_{\mathrm{NB}(r,p)}(t) &= \mathbb{E}\left[\exp\left(itY\right)\right] \\
&= \sum_{y=0}^{\infty} \exp(ity)\, \frac{\Gamma\left(y+r\right)}{\Gamma\left(y+1\right)\Gamma\left(r\right)}\, \left(1-p\right)^{y} p^{r} \\
&= \left(\frac{1}{p}\right)^{-r} \sum_{y=0}^{\infty} \frac{\Gamma\left(y+r\right)}{\Gamma\left(y+1\right)\Gamma\left(r\right)} \left(1-p\right)^{y} \exp(it)^{y} \\
&= \left(\frac{1}{p}\right)^{-r} \left[1-\left(1-p\right)\exp(it)\right]^{-r} \\
&= \left[\frac{1-\left(1-p\right)\exp(it)}{p}\right]^{-r} \equiv p^{r}\left[1-\left(1-p\right)\exp(it)\right]^{-r}.
\end{aligned} \tag{7}$$

Since the base of $\phi_{\mathrm{NB}(r,p)}(t)$ is independent of $r$, it follows that if $\{Y_i\}_{i=1}^{n} \sim$ i.i.d. $\mathrm{NB}\,(r, p)$, then

$$Z := \left(\sum_{i=1}^{n} Y_i\right) \sim \mathrm{NB}\left(nr, p\right). \tag{8}$$

Moreover, since the moment generating function of $Y \sim \mathrm{NB}\,(r, p)$ is given by

$$M_{\mathrm{NB}(r,p)}(t) = \left[\frac{1-\left(1-p\right)\exp(t)}{p}\right]^{-r}, \tag{9}$$

we have

$$
\begin{aligned}
M'_{\mathrm{NB}(r,p)}(t) &= \frac{d}{dt}\left[\frac{1-(1-p)\exp(t)}{p}\right]^{-r} \\
&= \left[\frac{1-(1-p)\exp(t)}{p}\right]^{-r-1}(-r)\,\frac{d}{dt}\left[\frac{1-(1-p)\exp(t)}{p}\right] \qquad (10)\\
&= \left[\frac{1-(1-p)\exp(t)}{p}\right]^{-r-1}\frac{1-p}{p}\,r\,\exp(t),\ \text{and} \\
M''_{\mathrm{NB}(r,p)}(t) &= \frac{d}{dt}M'_{\mathrm{NB}(r,p)}(t) \\
&= \frac{d}{dt}\left[\left[\frac{1-(1-p)\exp(t)}{p}\right]^{-r-1}\frac{1-p}{p}\,r\,\exp(t)\right] \\
&= \frac{1-p}{p}\,r\left[\left[\frac{1-(1-p)\exp(t)}{p}\right]^{-r-1}\exp(t)+\exp(t)\frac{d}{dt}\left[\frac{1-(1-p)\exp(t)}{p}\right]^{-r-1}\right] \\
&= \frac{1-p}{p}\,r\left[\left[\frac{1-(1-p)\exp(t)}{p}\right]^{-r-1}\exp(t)+\exp(t)\left[\frac{1-(1-p)\exp(t)}{p}\right]^{-r-2}\frac{1-p}{p}\,(r+1)\,\exp(t)\right].
\end{aligned}
$$

(11)

So the mean and variance of $Y \sim \mathrm{NB}\,(r,p)$ are given by

$$
\begin{aligned}
\mathbb{E}[Y] &= M'_{\mathrm{NB}(r,p)}(t)\Big|_{t=0} \\
&= \left[\frac{1-(1-p)\exp(t)}{p}\right]^{-r-1}\frac{1-p}{p}\,r\,\exp(t)\Bigg|_{t=0} \qquad (12)\\
&= \frac{1-p}{p}\,r,\ \text{and} \\
\mathbb{V}[Y] &= \left[M''_{\mathrm{NB}(r,p)}(t)\Big|_{t=0}\right]^2-\mathbb{E}^2[Y] \\
&= \frac{1-p}{p}\,r\left[\left[\frac{1-(1-p)\exp(t)}{p}\right]^{-r-1}\exp(t)+\exp(t)\left[\frac{1-(1-p)\exp(t)}{p}\right]^{-r-2}\frac{1-p}{p}\,(r+1)\,\exp(t)\right]\Bigg|_{t=0} \\
&\quad -\left(\frac{1-p}{p}\,r\right)^2 \\
&= \frac{1-p}{p}\,r\left[1+\frac{1-p}{p}\,(r+1)\right]-\left(\frac{1-p}{p}\,r\right)^2 \\
&= \frac{1-p}{p^2}\,r,
\end{aligned}
$$

(13)

respectively. Consequently, we have

$$
\mathbb{E}[Z] = \frac{1-p}{p}\,nr = n\mathbb{E}[Y],\ \text{and} \qquad (14)
$$

$$
\mathbb{V}[Z] = \frac{1-p}{p^2}\,nr = n\mathbb{V}[Y]. \qquad (15)
$$

### 2 Supplementary Figures

#### 2.1 Histograms

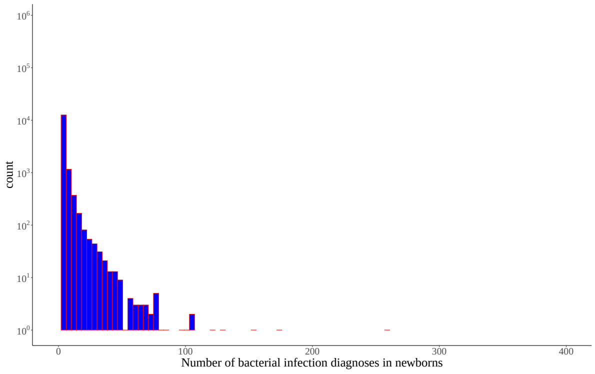


(a) Bacterial infections.

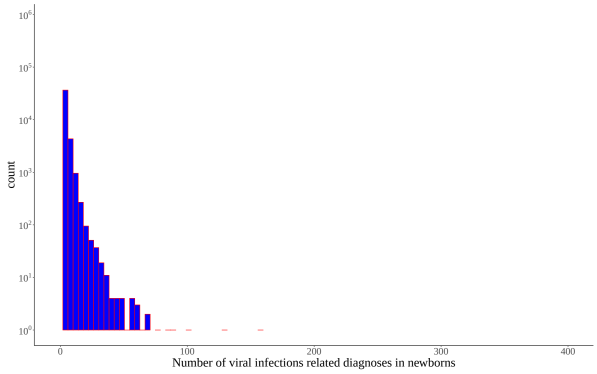


(b) Viral infections.

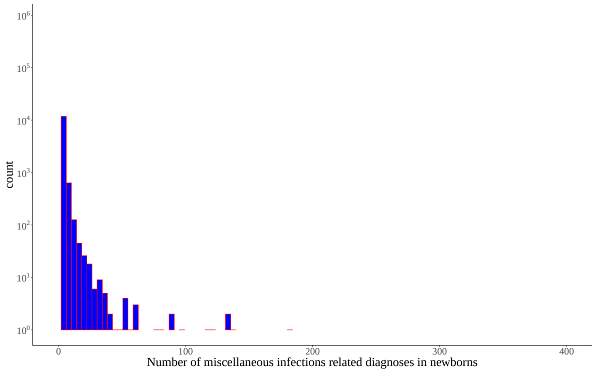


(c) Miscellaneous infections.

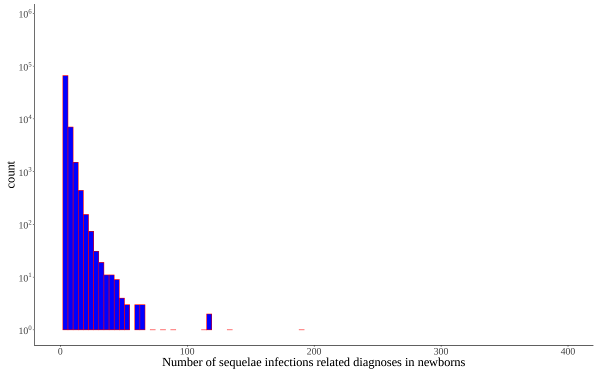


(d) Sequelae infections.

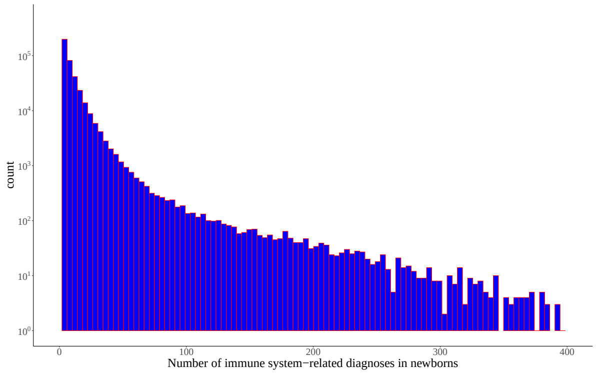


(e) Immune disorders.

Figure S7: Histograms of counts of phenotypes

### 3 ICD9 and 10 codes

See raw text files.

### References


[1] Alan Agresti. *Foundations of linear and generalized linear models*. John Wiley & Sons, 2015.

[2] IBM Watson Health. *IBM MarketScan Research Databases for life sciences researchers*. 2019.

[9] Rainer Winkelmann. *Econometric analysis of count data, 5th edition*. Springer Science & Business Media, 2008.

[10] World Health Organization. *The ICD-10 classification of mental and behavioural disorders: clinical descriptions and diagnostic guidelines*. World Health Organization, 1992.
